# Using AI to support rapid qualitative data analysis of survey and interview data in public health: a proof-of-concept study

**DOI:** 10.64898/2026.06.19.26356035

**Authors:** Tom White, Riinu Pae, Carina Hörst, Avelie Stuart, Rachel Abbey, Elena Skryabina, Samantha Brooks, Paulina Bondaronek, Lorenzo Cattarino

## Abstract

**Background:** Large language models (LLMs) are increasingly used to support a growing range of analytical and operational tasks in public health, and show promise for assisting qualitative text analysis at scale. While they have demonstrated utility in structured natural language processing tasks, their role in more interpretive approaches such as thematic analysis remains less clear. Thematic analysis requires careful, often time-intensive engagement with qualitative data, which can be challenging when datasets are large or when policy teams must deliver rapid insights. This study examines whether a pragmatic LLM-assisted workflow can support early-stage thematic analysis of large public health datasets in a way that is systematic, transparent, and compatible with human-led analytical oversight.

**Methods:** We developed and evaluated a proof-of-concept workflow for LLM-assisted semantic coding and theme identification in a public health case study involving qualitative survey and debrief data. The LLM generated codes and themes using a consistent prompting structure. We compared those themes with those produced through human-only thematic analysis and with outputs from a topic-modelling based approach. A convergence coding matrix was used to classify alignment as agreement, complementarity, disagreement or silence. To evaluate the reliability of the LLM’s theme classification, we manually labelled 500 codes and calculated accuracy metrics.

**Results:** The LLM-generated themes showed broad alignment with the human analysis at the level of higher-order themes, with agreement observed for 73% of manual themes. Complementary LLM themes were found for a further 18%, while only one subtheme showed dissonance and a small number (8.3%) had no clear match. Compared with the topic modelling approach, the LLM produced a broader and more detailed thematic structure. In the theme assignment task, the LLM achieved an overall F1 score of 0.68, with individual theme scores between 0.34 and 0.80, indicating moderate consistency with humans and stronger performance for clearly defined themes.

**Conclusions:** These findings suggest that LLM-assisted thematic analysis has promise as a pragmatic proof-of-concept approach for rapid, higher-level qualitative sensemaking in public health, particularly when datasets are large and timely insight is needed. Its use should remain bounded to surface-level exploratory analysis and embedded within human-led workflows.

## Background

Recent advancements in generative artificial intelligence, particularly the emergence of large language models (LLMs), offer promising new avenues for automating and enhancing how we analyse text data at scale. These models, trained on vast amounts of data, possess the ability to recognise patterns in and generate human-like text, making them well-suited for analysing large volumes of qualitative data. LLMs have been applied across a range of domains to perform a wide array of text-processing tasks including text annotation (Alizadeh et al., 2025), summarisation (Aali et al., 2024) and sentiment analysis (Miah et al., 2024) – current applications and future challenges have been covered in detail in Kaddour et al. (2023). In public health, LLMs have also been used for classification and extraction tasks, such as investigating infectious disease outbreaks from online reviews (Harris et al., 2025) and understanding sentiment towards vaccination from social media (Espinosa & Salathe, 2024). Such Natural Language Understanding tasks (Wong et al., 2023) can be thought of as a classification exercise and can be validated using established methods for measuring accuracy.

Much less is known, however, about the role LLMs might play in qualitative research practices that rely on judgement, transparency and careful engagement with data. Thematic analysis remains one of the most widely used approaches for examining open-ended survey responses and interview transcripts, particularly within health research (Richardson, 2009; Braun, 2014; Saunders, 2023) and the implementation of widescale interventions (Turner et al., 2022). While reflexive thematic analysis emphasises prolonged, thoughtful engagement with data and the active, interpretive role of the researcher (Braun & Clarke 2008; 2019), other forms of thematic analysis focus on mapping the data onto pre-existing coding frameworks or looking for data prevalence (Vaka, 2021). Regardless of the specific qualitative approach used, challenges often arise when researchers must analyse very large datasets or produce findings rapidly. In these contexts, machine-assisted analysis can help by supporting researchers when the scale or urgency of qualitative data exceeds what human teams can reasonably analyse during fast-moving situations such as public health emergencies. This reflects common critiques of thematic analysis, particularly the time and labour required to manually code large datasets. These pressures have prompted growing interest in whether LLMs might support or accelerate qualitative analysis, particularly in contexts such as outbreak management and pandemic response.

Some attempts have been made to use open-source, publicly available LLMs for thematic analysis (Dai et al., 2023; De Paoli, 2024; Drápal et al., 2023; Mathis et al., 2024). These studies suggest that LLMs can generate coherent themes from small interview datasets, but important questions remain about their validity, scalability, reproducibility, and reliability when applied to larger public health datasets. Additionally, the opaque nature of model reasoning continues to raise concerns about transparency and trust (Liao, 2024) - factors that are essential when qualitative findings inform operational decisions, especially if at a national scale. Understanding the limitations and trade-offs of LLM-assisted analysis, including the balance between automation and human judgement, is therefore crucial before these tools can be meaningfully integrated into public health practice.

In this paper, we position LLM-assisted thematic analysis not as a replacement for human-led qualitative inquiry, but as a pragmatic proof-of-concept for rapid analysis in public health settings where teams must make sense of large volumes of textual data under time and resource constraints. Our focus is therefore on whether such an approach can support timely, higher-level pattern identification and transparent human review, rather than on whether it can reproduce the full interpretive depth of human-led thematic analysis. Our objectives are:

1. To assess how closely LLM-generated outputs align with human-generated themes in a proof-of-concept public health case study.
2. To examine the transparency and practical usefulness of the workflow by mapping codes to themes and evaluating classification accuracy under human review.

## Methods

### Covid-19 Case Study

To enable a robust comparison between our LLM-assisted methods, developed specifically for this study, and established qualitative methods, we selected a case study that was being examined concurrently by an experienced research team using traditional thematic analysis techniques (Skryabina, In prep). The aim of the study was to better understand the sources of organisational learning during a major public health emergency. An advantage of this case study was its use of multiple data sources, which allowed us to examine how our approach functioned across different kinds of qualitative data formats. The dataset comprised surveys and debriefs from public health professionals involved in the UK COVID-19 response between February and September 2020, covering topics such as coordination, information flows, and broader reflections collected at multiple points throughout the pandemic. These materials offered rich insight into operational experiences and informed the aims of the original analysis, which sought to better understand the sources of organisational learning during a major public health response. Working with a dataset undergoing traditional expert analysis allowed us to directly assess where LLM-generated insights aligned with, diverged from, or complemented those derived through conventional qualitative methods. It also reflects a real public health context in which large volumes of qualitative data may need to be analysed quickly to support organisational learning during or after a major incident.

### Data

We utilised two primary qualitative data sources to align with the original analysis: open-ended staff survey responses and semi-structured debrief interviews. Because these sources differed in structure and depth, each required tailored preprocessing to enable fair comparison in the subsequent analysis. All data were pre-processed in Python, to remove missing values and ensure they were in a tabular format appropriate for the downstream analysis by an LLM. Text from debriefs had to be extracted from raw word documents and converted to tabular data, where each row was the answer to an interview question.

The surveys were completed by personnel involved in the Covid-19 response across professional grades in Public Health England, conducted across three operational phases: contain (3^rd^ – 17^th^ April 2020), delay (15^th^ – 29^th^ May) and post-lockdown (20^th^ July – 4^th^ August 2020). Although the surveys included several directed questions, our analysis focused on the open-ended items that invited broader reflection: (a) key learnings that should be reinforced in future responses, (b) elements that could be improved and reasons why, and (c) any additional comments. Across all phases these open-ended questions yielded 1,142 responses from 682 participants. Because staffing changed across operational phases, the surveys do not represent repeated measures, and different personnel may have contributed responses at each wave.

Semi-structured debrief interviews were conducted in June – September 2020 with senior staff and tactical leads, structured around seven thematic domains (Your Role; Preparation and Expectations; Governance and Decision Making; Information Flows; Partnership Working and Coordination; Resource and Wellbeing; Communication). For each domain, participants were asked (a) what worked well, (b) what did not work well, (c) what they would do differently and (d) for any additional comments. These debriefs produced 842 responses from 54 debriefs across domains and questions, which were all treated as part of one dataset for our analysis.

### Data analysis

#### Model

Previous work carried out by Harris and colleagues developed an internal LLM API on UKHSA High Performance Computing resources and evaluated a range of models across several public health classification and extraction tasks (2025). We used this API and the model which they had shown to consistently perform best across most tasks, Llama-3.3-70B-Instruct (AI@Meta, 2024).

#### Analytical method

We designed this workflow as a pragmatic proof-of-concept rather than as a replacement for human-led thematic analysis. The method was intentionally scoped to support rapid, semantic-level pattern detection in a public health context where timely overview may be more valuable than fine-grained interpretive depth.

We developed an analytical approach for conducting thematic analysis using Large Language Models, following Braun and Clarke’s (Braun, 2008) established process for thematic analysis. Their approach for thematic analysis comprises 6 steps:

1. Familiarising with data
2. Generating initial codes
3. Creating themes
4. Reviewing themes
5. Defining and naming themes
6. Producing the report

Because LLM-based processing is automated, we drew on this process pragmatically. Specifically, we adapted stages 2 and 3 by using the LLM to support coding and theme development, as it is well suited to summarising and organising discrete units of meaning. Themes were conceptualised as coherent “patterns of shared meaning” (e.g. ideas, concepts, experiences) “underpinned by a central organising concept” (Braun & Clarke, 2019, p. 589). The LLM-produced themes and subthemes were then reviewed by a human before being finalised.

#### Prompt engineering

All prompts followed a consistent structure. The system section of the prompt defined the LLM’s role (an expert qualitative researcher), provided context about the dataset (reflections on the Covid-19 response), and stated the overall analytical goal (identifying learning points via thematic analysis). The user section of the prompt for the coding stage specified the task and constraints for the model, including definitions of codes (semantic features of the text), the required output format, and the text to be analysed. Finally, an assistant header within the prompt indicated the point at which the model should begin its response.

Theme identification followed the same prompting structure but replaced the response text with the list of codes. The user section additionally included a definition of themes as ‘patterns of meaning organised around a central idea’ and specified the desired structure of the thematic output.

#### Process

The main analysis proceeded in two stages. First, each individual data element (survey response or debrief segment) was submitted to the LLM as part of a structured prompt designed to elicit a set of potential codes (step 1, Figure 1). Next, we aggregated all resulting codes and prompted the LLM again to identify overarching themes and subthemes (step 2, Figure 1). As a result of the model’s restricted processing capacity (its *context window* was restricted to the equivalent of about 8,000 words), we split the dataset into smaller sections. For each section we provided the full list of codes that the model had generated from that subset and asked it to propose possible themes. A final consolidation prompt combined the chunk-level themes into a unified set. This ensured that the thematic structure was derived from the entire dataset while remaining within the model’s technical constraints.

**Figure 1.**
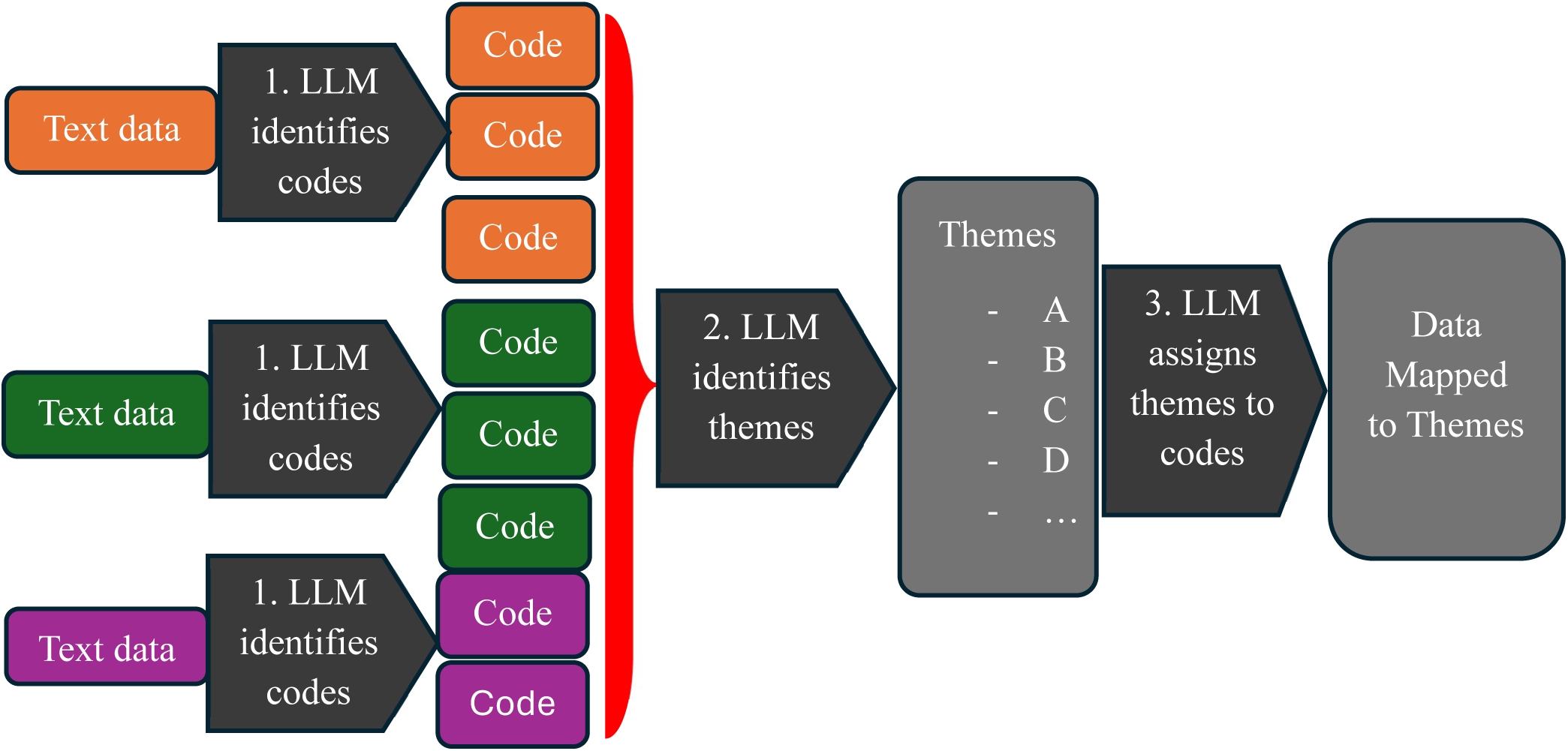
Flow diagram of analytical process. The LLM first identifies semantic codes within the text (1), then identifies themes within these codes (2), and finally assigns themes to codes (3).

We edited the LLM’s final list of themes and subthemes to improve coherency and clearness. This included removing subthemes that don’t provide further information and rewording of themes for clarity. This aligns with Braun and Clarke’s steps 4 and 5 (reviewing and defining themes).

Finally, to understand which responses drove individual LLM-generated themes and examine wider explainability of LLM outputs, we first needed to determine which codes contributed to each theme (step 3, Figure 1). We prompted the LLM to classify each previously generated code into one of the identified themes, instructing the model to assign a code to “No suitable theme” when none of the themes were a close semantic match, consistent with human-led thematic analysis where not all generated codes align with the final themes. This produced a mapping from codes to themes that could be used to quantify their distribution and prevalence, and provide greater clarity and transparency into how the themes were generated by the LLM.

### Evaluation of LLM-based approach

To assess the alignment between the LLM-generated themes and those produced through human-only thematic analysis, we conducted a structured triangulation exercise (Farmer, 2006). This process is based on using a convergence coding matrix to classify the relationship between each pair of themes as one of four categories: Agreement (conceptual alignment); Complementarity (shared meaning with some differences in emphasis); Dissonance (substantive disagreement); or Silence (present in only one analysis). Three researchers (TW, AS, RP) independently applied these classifications before discussing and resolving any differences. This process enabled a systematic assessment of overlap and divergence between human and LLM produced themes.

To contextualise the performance of our LLM-based approach further, we also conducted topic modelling (a machine learning approach) with qualitative interpretation of the findings. We followed the approach introduced as Machine-Assisted Topic Analysis (Towler, 2023). A machine learning model is first used to organise large-scale data into topics, making it manageable, with the number of topics selected to optimise perplexity (how well the model classifies unseen data) and topic diversity (how distinct topics are from each other). Qualitative researchers then take over and conduct in-depth analysis on 20 representative responses from each generated topic, identifying themes and assigning a hierarchy of subthemes as required. These themes were compared with the human themes using the same four triangulation categories.

Finally, to assess the reliability of the codes-to-themes classification step (step 3 in Figure 1), we manually labelled a subset of 500 randomly selected codes and compared these human-generated labels with the LLM’s assignments to calculate accuracy metrics. We selected a sample of 500 codes because this provided a sufficiently large set to yield stable accuracy estimates while remaining feasible for manual annotation. The manual dataset was created through a structured, multi-stage process designed to ensure consistency and to establish a reference point for evaluating model performance. First, all four researchers (TW, LC, CB, SF) independently labelled the same set of 50 codes with themes. We then compared our decisions, discussed points of disagreement, and collaboratively developed labelling guidelines to support a consistent approach. This stage also provided insight into the natural variation that occurs even when humans follow the same instructions. After the guidelines were agreed, the remaining 450 codes were divided evenly across the research team, with each code being labelled by a single researcher. Any cases that raised uncertainty were brought back to the group for discussion and consensus. This procedure allowed us to construct a reliable human-annotated dataset against which to benchmark the model’s ability to assign codes to the appropriate themes.

Using the human-verified labels as the reference standard, we calculated theme-level and overall precision, recall and F1-scores. Precision reflects the proportion of the LLM’s assigned themes that were correct, recall indicates the proportion of true human-assigned themes that the LLM successfully retrieved, and the F1-score represents the harmonic mean of precision and recall, providing a single balanced measure of performance. All metrics are between zero (worst possible performance) and one (perfect performance).

## Results

### Themes

The themes generated by our LLM-assisted thematic analysis were compared against those reported in (Skryabina, In prep), which served as the reference point for this study. Using our analysis framework, the model produced a coherent set of themes that captured the recurring patterns within the dataset. The major themes identified through the LLM-assisted process together with those identified through traditional qualitative analysis and through the ML + human approach are shown in table 1.

**Table 1.** Themes produced from the three analysis techniques: traditional, LLM and ML + human. Themes do not align across rows.

| Traditional Themes |  | LLM Themes |  | ML + human |
| --- | --- | --- | --- | --- |
| 1. Effective Communication<br>- Importance of meetings and briefings<br>- Importance of regular updates in a rapidly evolving situation<br>- Importance of simple systematic communication | 7. Welfare and Wellbeing<br>- Importance of wellbeing support<br>- Need for recognition<br>- Workload and job demands<br>- Stress<br>- Workplace relationships<br>- Positive impacts on staff | 1. Collaborative Approach<br>- Interagency collaboration and partnership<br>- Team Dynamics and coordination | 6. Learning and Improvement<br>- Lessons Learned and areas for improvement<br>- Reflection and appreciation | 1. Communication<br>- Need to improve communication<br>- Developing and communicating guidance<br>- Frequent meetings and updates |
| 2. Effective Coordination and collaboration<br>- Importance of good coordination within PHE<br>- Importance of cross-organisation coordination<br>- Duplication of effort<br>- Working in silos<br>- Allocation of work | 8. Information and Data Sharing<br>- Information flow<br>- Sharing data | 2. Infrastructure and Resource Challenges<br>- Resource allocation and management<br>- Staffing and resource constraints<br>- Technology, infrastructure and systems limitations | 7. Preparedness and planning<br>- Scenario planning, exercise, and drills<br>- Capacity building and succession planning | 2. Pandemic Preparedness and Planning |
| 3. Capacity<br>- Importance of sufficient capacity<br>- Cell set-up<br>- Rota management<br>- Importance of continuity<br>- Importance of trained staff<br>- Importance of staff commitment<br>- Recruitment | 9. Technology and Resources<br>- Challenges with technology<br>- Benefits of technology<br>- Challenges of lack of resources | 3. Effective Communication<br>- Communication challenges and difficulties<br>- Importance of communication in the response effort<br>- Clear guidance and daily updates<br>- Centralised information and core briefing | 8. External Factors and Influences<br>- Government policies and guidance<br>- Public perception and messaging<br>- Media coverage and reputation management<br>- Public interaction and engagement. | 3. Collaboration<br>- Challenges establishing PHE's role within the response<br>- Inter-cell collaboration<br>- Within-cell collaboration<br>- Strength of the working group and how they collaborate |
| 4. Leadership and Governance<br>- Poor leadership<br>- Good leadership<br>- Managing expectations<br>- Decision-making processes | 10. Working Location<br>- Importance of shared working locations with colleagues<br>- Benefits of working from home | 4. Leadership and Governance<br>- Leadership and decision-making<br>- Governance and accountability<br>- Clear role definitions and accountability<br>- Leadership critique and feedback | 9. Process Improvement and Development<br>- Process efficiency and improvement<br>- Technology and infrastructure development<br>- Information management and data analysis<br>- Organisational improvement and adaptation | 4. Resource Management<br>- Resources adequate but needed more support<br>- Challenges regarding sustainability of workforce<br>- Need to utilise training and expertise<br>- Challenges onboarding the response |
| 5. Planning and Preparedness<br>- Importance of preparedness<br>- Need to be proactive not reactive<br>- Importance of adaptability and flexibility<br>- Addressing inequalities | 11. Organisational Learning<br>- Importance of learning for future response<br>- Lack of time for reflecting | 5. Staff Wellbeing and Support<br>- Workload management and burnout<br>- Recognition of staff contributions<br>- Emotional wellbeing and support | 10. Organisational Culture and Values<br>- Organisational culture and structure<br>- Autonomy and decision-making<br>- Organisational values and principles | 5. Cell Management |
| 6. Role Clarity<br>- Personal role clarity<br>- Cell remits<br>- Clarifying role of PHE to others<br>- Clear processes in place |  |  |  | 6. Operational challenges due to lack of time/resource |
|  |  |  |  | 7. Data handling and Information Sharing |

### Triangulation

The formal triangulation process showed that the LLM-generated themes and subthemes were broadly similar to those produced by the human-led analysis (Figure 2). 73% of themes (8/11) and 29% of subthemes (12/42) from the human analysis were in agreement with LLM-generated themes and subthemes. These included, for example the theme “Welfare and Wellbeing” (human generated), which agreed with “Staff Wellbeing and Support” (LLM generated). 18% of themes (2/11) and 55% of subthemes (23/42) from the human-led analysis had at most complementary counterparts in the LLM-generated sets. For example, the human-generated theme “Technology and Resources” was complementary to the LLM-theme “Infrastructure and Resource Challenges”. None of the 12 themes and just one of the 42 subthemes (2.4%) from the human-led analysis were in disagreement with the LLM-generated themes and subthemes. The human-led analysis subtheme “Lack of time for reflecting” was in contradiction with the LLM-generated subtheme “Reflection and appreciation”. 8.3% of the themes (1/12) and 14% of subthemes (6/42) from the human-led analysis did not correspond either positively or negatively with any of the LLM-generated themes and subthemes, and so were assigned the “silence” label. These included for example the theme on “Working Location” and its underlying subthemes. Figure 2 shows these results together with how they compare against the ML + human approach. Taken together these results suggest that the LLM-assisted workflow is more effective for rapid identification of broad thematic structure than for reproducing finer-grained qualitative distinctions.

**Figure 2.**
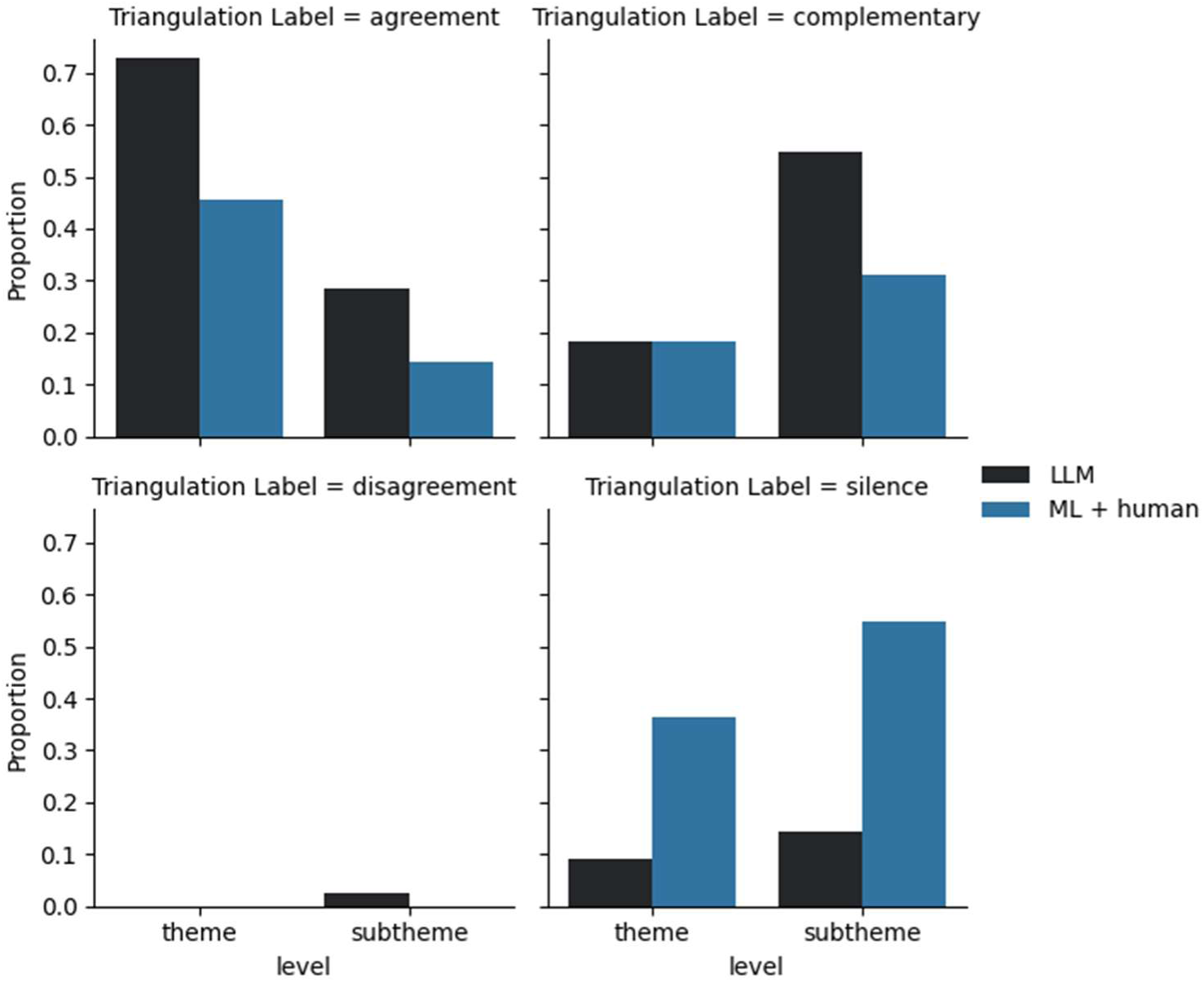
Bar charts showing the proportion of themes and subthemes in each of the triangulation categories when comparing the LLM (black) and ML + human (blue) outputs with the manually produced themes.

**Figure 3.**
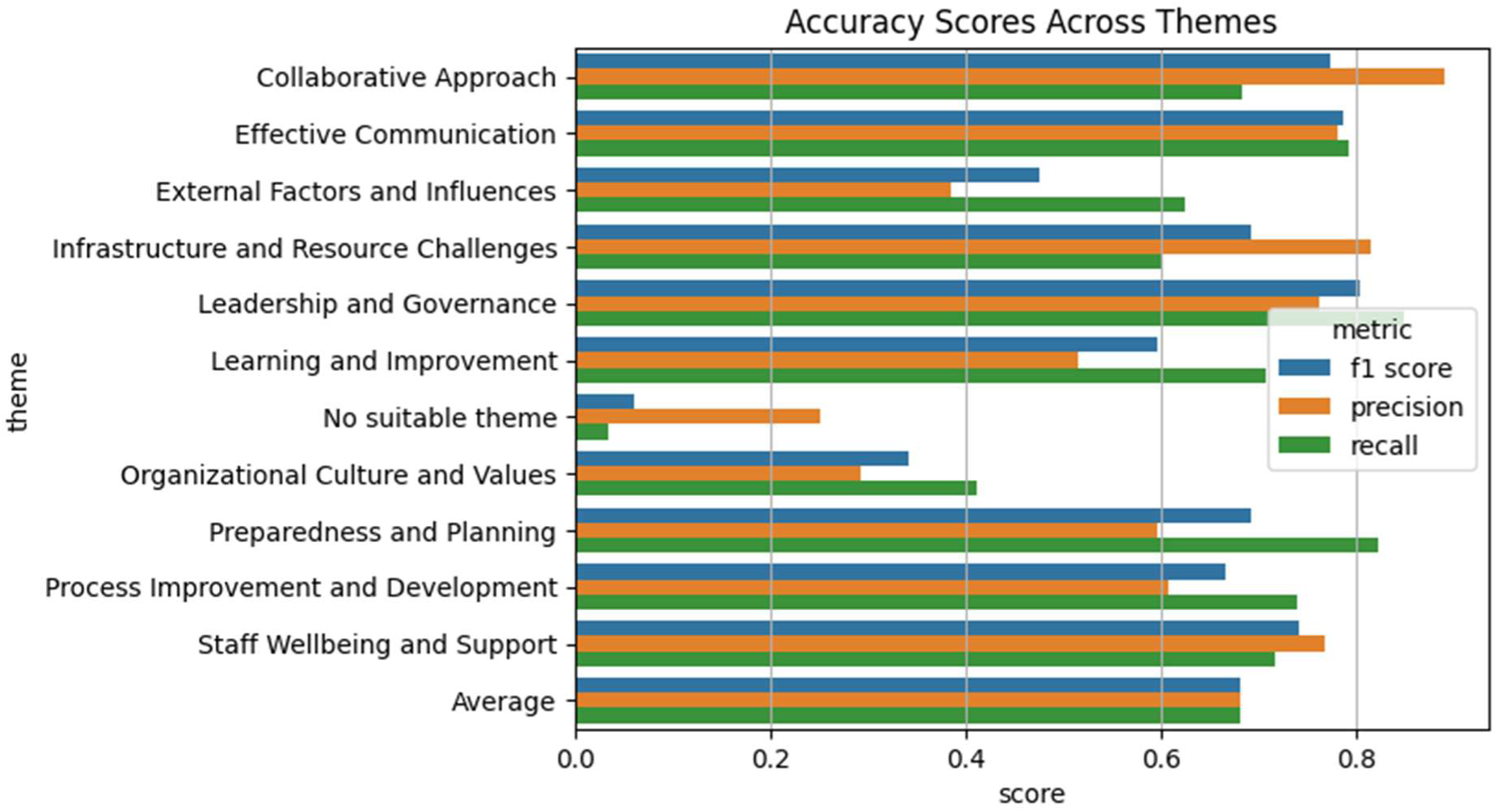
Code classification performance metrics across themes.

### Mapping Accuracy

To evaluate the accuracy with which the LLM can assign themes to codes, we compared its outputs against a manually labelled subset of 500 codes. Using the human-verified labels as the reference standard, we calculated theme-level and overall precision, recall and F1-scores. The model achieved an overall F1-score of 0.682, with individual theme scores ranging from 0.341 (for Organisational Culture and Values) to 0.804 (for Leadership and Governance), reflecting variability in agreement between the LLM and human coders across themes.

## Discussion

This study was a pragmatic proof-of-concept exploring whether LLM-assisted analysis can support rapid, higher-level qualitative sensemaking in public health. Rather than evaluating LLMs as substitutes for human qualitative researchers, we examined whether a structured, transparent workflow could help identify broad patterns in large text datasets in contexts where time pressures and data volume challenge traditional approaches. To assess this, we compared the outputs of the LLM-assisted process with both human-only thematic analysis and a topic-modelling-based method. The discussion therefore focuses on the specific value of this approach as a rapid, exploratory, human-supervised tool in public health, as well as its limitations for more detailed interpretive work.

Our findings demonstrate that LLM-assisted thematic analysis can produce outputs that broadly align with human-generated themes, with particularly strong correspondence at the level of higher-order thematic structure. The triangulation exercise showed that most human-generated themes had clear counterparts in the LLM analysis, and where differences emerged, they were typically complementary rather than contradictory. Notably, this convergence was far stronger at the level of overarching themes than at the level of subthemes. Only 29% of subthemes developed through human-led analysis were judged to be in agreement with the LLM outputs, suggesting that the model captured broader patterns more consistently than the more detailed distinctions identified by human analysts. These results indicate that LLM-assisted thematic analysis is well suited to higher-level exploratory work and can function as a breadth-oriented analytical tool in the sense described by Weller’s Breadth-and-Depth approach, supporting the systematic mapping of large corpora to highlight patterns and guide subsequent in-depth, human-led analysis (Davidson, 2019). Future research should explore how best to combine these approaches in practice, particularly how more detailed, interpretive analysis can build on an initial LLM-assisted, high-level mapping without requiring researchers to restart from raw data. This could include comparing different sequences of human–LLM interaction (e.g. LLM-led mapping followed by human interpretation, human immersion followed by LLM-supported synthesis, or concurrent approaches), as well as examining how reflexive practices can be meaningfully incorporated into such workflows. Further work is also needed to identify the types of themes or concepts that LLMs handle less well, particularly more abstract, context-dependent constructs, in order to clarify when their use may be less appropriate. Insights from emerging rapid qualitative methods, such as RREAL (Rapid Research Evaluation and Appraisal Lab) approaches, may offer a useful starting point for developing such combined workflows. This stands in contrast to the triangulation comparing the topic-modelling approach, which captured only a subset of the manual themes and struggled to represent the more nuanced patterns in the data.

The mapping-accuracy results reinforce the picture: although the LLM did not perfectly reproduce the human decisions, its moderate overall F1-score indicates that it was generally able to assign codes to appropriate thematic categories, with stronger performance in clearly defined domains. However, performance varied widely across themes (F1-scores ranging from 0.341 to 0.804), suggesting uneven reliability rather than uniform accuracy. The themes with weaker results tended to be the ones where the boundaries were fuzzy or where meaning depended on background knowledge that the model didn’t have, such as the theme “Organisational Culture and Values”. This pattern suggests that LLM-assisted theme assignment is more dependable for concrete, well-bounded topics than for abstract or ambiguous domains where interpretation relies on tacit understanding.

Importantly, complete convergence is not a realistic benchmark; qualitative analysis is inherently subjective and independent human-led analyses seldom achieve perfect alignment. From this perspective, the areas of divergence between the LLM and manual interpretations are better understood as reflecting the natural variation that arises whenever different analysts, human or machine, engage with the same dataset, rather than signalling a deficiency unique to the model. At the same time, the observed variation across themes underscores the need for cautious interpretation of aggregate performance metrics and highlighting the need for careful interpretation in more conceptually complex domains. Together, these findings suggest that LLMs can support systematic pattern identification in large qualitative datasets in ways that surpass traditional automated methods, while still requiring human oversight to ensure interpretive fidelity. These comparisons should nonetheless be interpreted cautiously.

While these findings demonstrate the value of LLMs for supporting large-scale qualitative analysis, they also clarify the conditions under which such tools are most appropriately used in practice. Recent commentary has highlighted strong objections to the use of generative AI in reflexive qualitative methodologies. Jowsey and colleagues (2025), for example, argue that reflexive thematic analysis is fundamentally a human, interpretive practice and therefore “not methodologically congruent” with the use of LLMs, also noting social and environmental justice concerns. However, this position is not uncontested. A growing body of work advocates for more pragmatic, “augmented” or human-in-the-loop approaches, which position LLMs as analytical aids rather than replacements for interpretation (Nguyen-Trung, 2026). These perspectives align closely with the approach taken in this study, where LLMs are used to support semantic, surface-level pattern identification within a broader human-led analytical process.

Within such workflows, careful consideration is needed to ensure that LLM outputs support rather than constrain analysis. For example, model-generated themes may unintentionally anchor researchers’ thinking or encourage over-reliance in time-pressured settings, limiting deeper engagement with the data. To mitigate these risks, LLMs should be used as analytical assistants to inform, rather than determine, interpretive decisions, with researchers actively interrogating and refining outputs. This is particularly important in contexts such as public health, where recent evidence suggests that LLM-generated qualitative outputs may be sensitive to demographic framing, raising concerns around representational bias and fairness (Bondaronek et al., 2026). Establishing practical guidance on how to integrate LLM-assisted insights into qualitative workflows will therefore be essential to ensure that their efficiency benefits are realised without compromising analytical rigour or equity considerations.

Establishing clear guidance on when and how to incorporate LLM-assisted insights will be essential to ensure that the benefits of these tools are realised without compromising the rigour and integrity of qualitative research. In practical terms, the workflow may be most valuable as a rapid analysis layer within public health intelligence and evaluation processes, helping teams to triage large qualitative datasets, surface emerging issues, and prioritise areas for deeper human-led analysis. At the same time, LLMs offer a distinctive advantage in their capacity to rapidly surface emerging themes and subthemes from qualitative data, making them particularly useful for informing decision making in fast-moving situations such as major incident responses, where timely insights can support initial sensemaking and prioritisation.

## Conclusions

This study provides proof-of-concept evidence that large language models may be useful for rapid, higher-level qualitative analysis of large text datasets in public health. The findings suggest that LLM-assisted workflows can support early-stage, semantic-level pattern identification and help reduce the burden of analysing large volumes of open-ended data, particularly in time-pressured or resource-constrained settings. However, the approach appears better suited to exploratory, surface-level sensemaking than to reproducing finer-grained interpretive distinctions characteristic of human-led qualitative analysis. It should therefore not be understood as a replacement for human-led thematic analysis, but as a bounded, pragmatic tool that can augment human analytical capacity when used within transparent, human-supervised workflows. Overall, the value of this approach lies in its potential to support timely public health insight generation while preserving a central role for human judgement, reflexivity and interpretive oversight.

## Data Availability

All data produced in the present study are available upon reasonable request to the authors.

## Declarations

### Ethics approval and consent to participate

Not applicable.

### Consent for Publication

Not applicable.

### Availability of data and materials

The datasets generated and/or analysed during the current study are not publicly available due to the inclusion of person-level staff feedback and the associated confidentiality and privacy considerations, but are available from the corresponding author on reasonable request.

### Authors’ contributions

TW prompted the large language model and interpreted the outputs together with LC, ES and SB. The ML + human analysis was carried out by AS and TW, with expert guidance from PB. The triangulation evaluation was completed by AS, RP and TW. Manual labelling of codes with themes was carried out by LC and TW. Writing the manuscript was lead by TW with key contributions from RP, CH, AS, RA, RA, PB, ES, and LC. All authors read and approved the final manuscript.

## Acknowledgements

We acknowledge the work of Charlotte Belsey and Sophie Ferguson, who assisted with manually labelling codes with themes to assess the accuracy of the mapping step.

## Appendices

### Prompts

The below are the text prompts we used to query the model at different stages of the analytical process.

### Identify codes

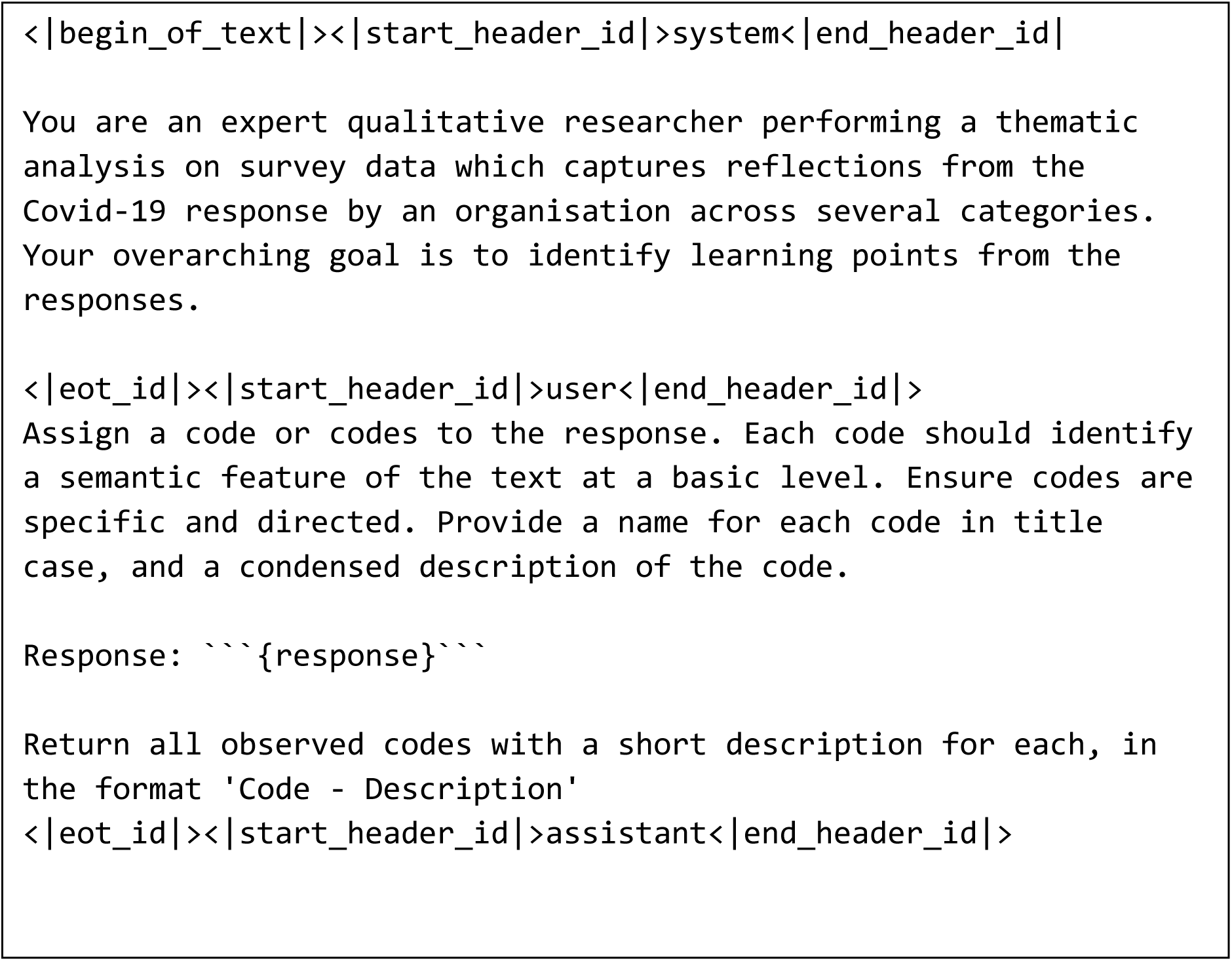

Where {response} was replaced with each survey response or interview answer in turn to identify themes within them.

### Identify Themes

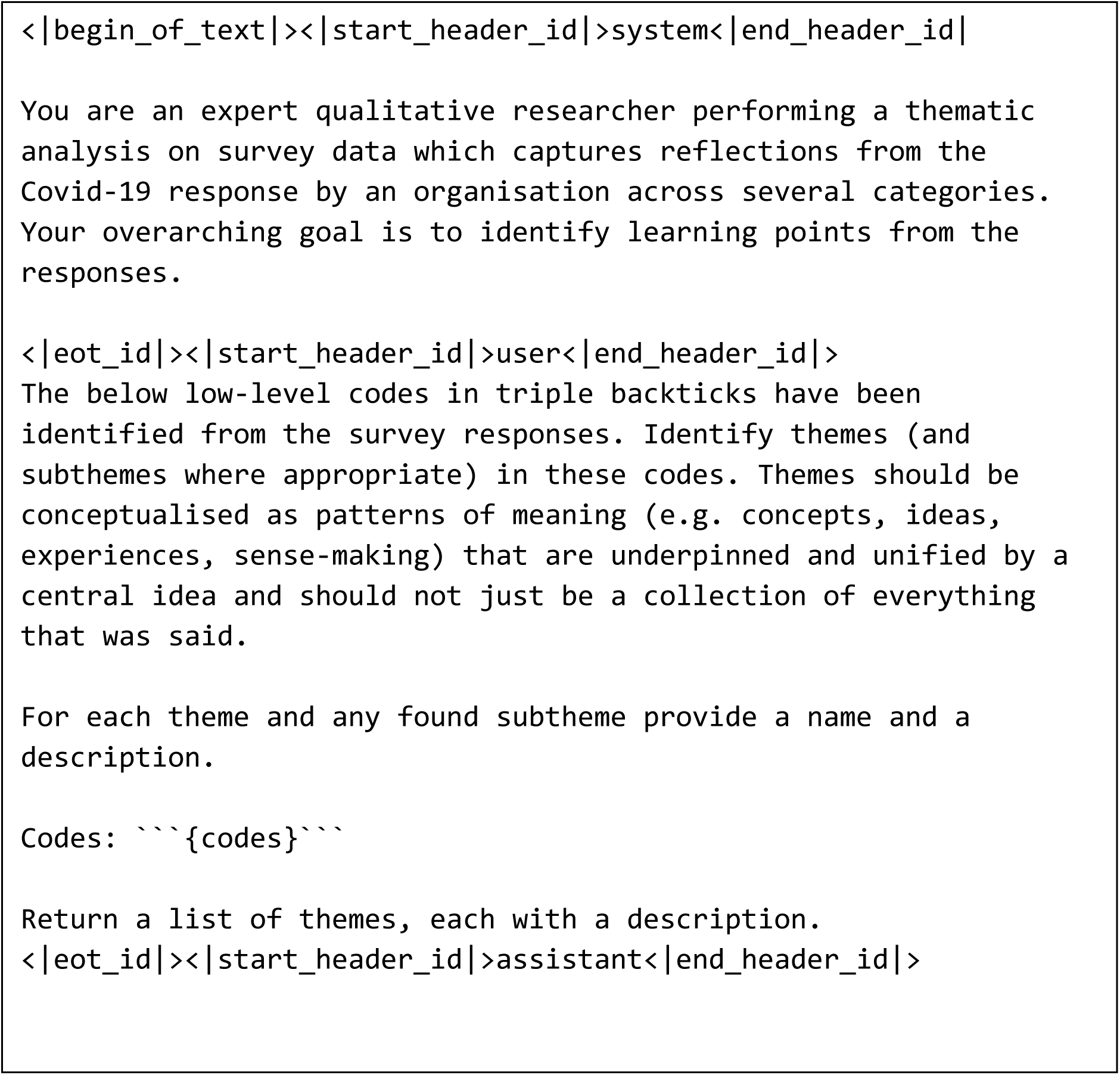

Where {codes} was replaced by the list of codes within that chunk to identify themes and subthemes.

### Consolidate chunks

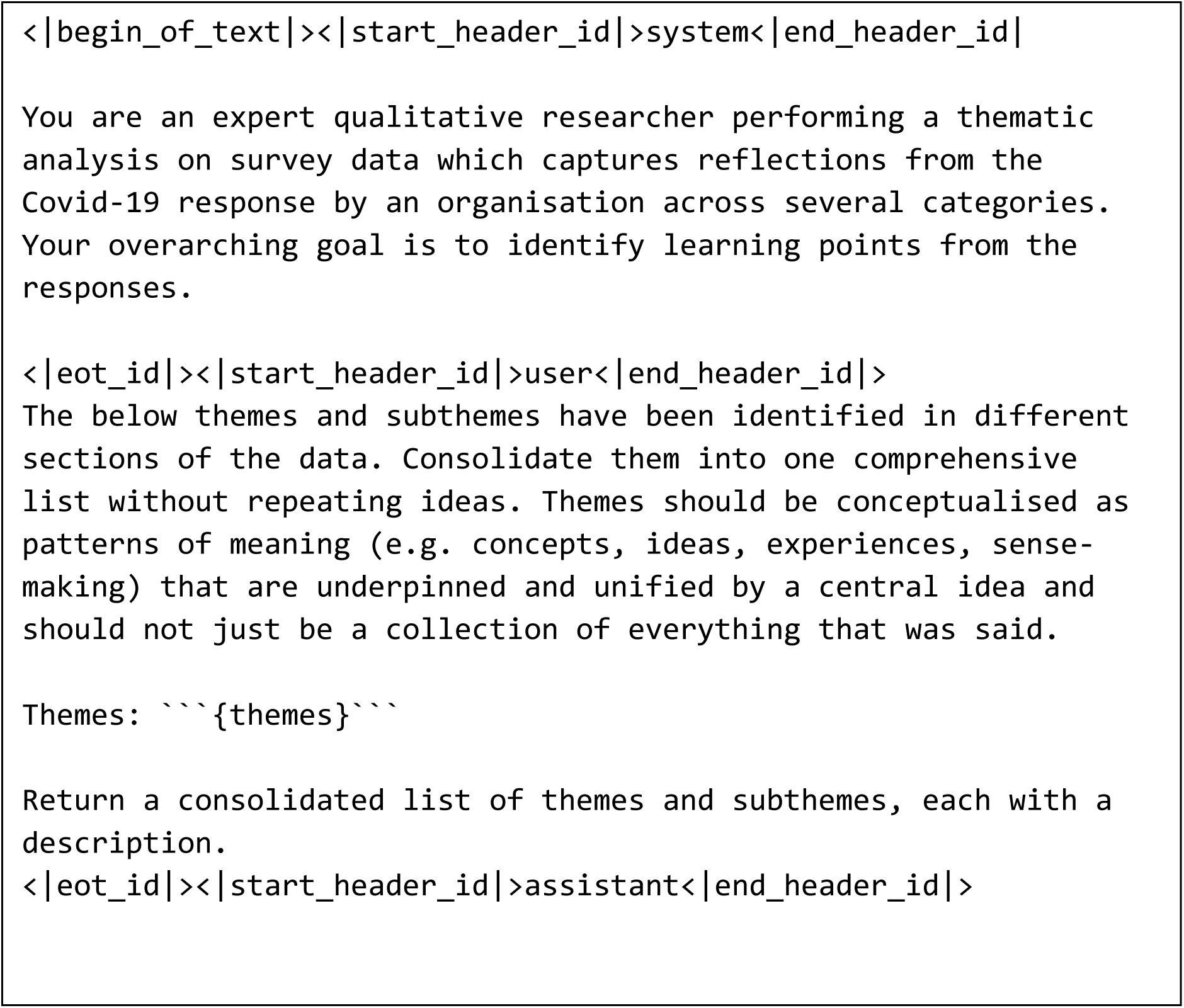

where {themes} is replaced with a list of all the themes and subthemes found in all of the chunks.

### Assign codes to themes

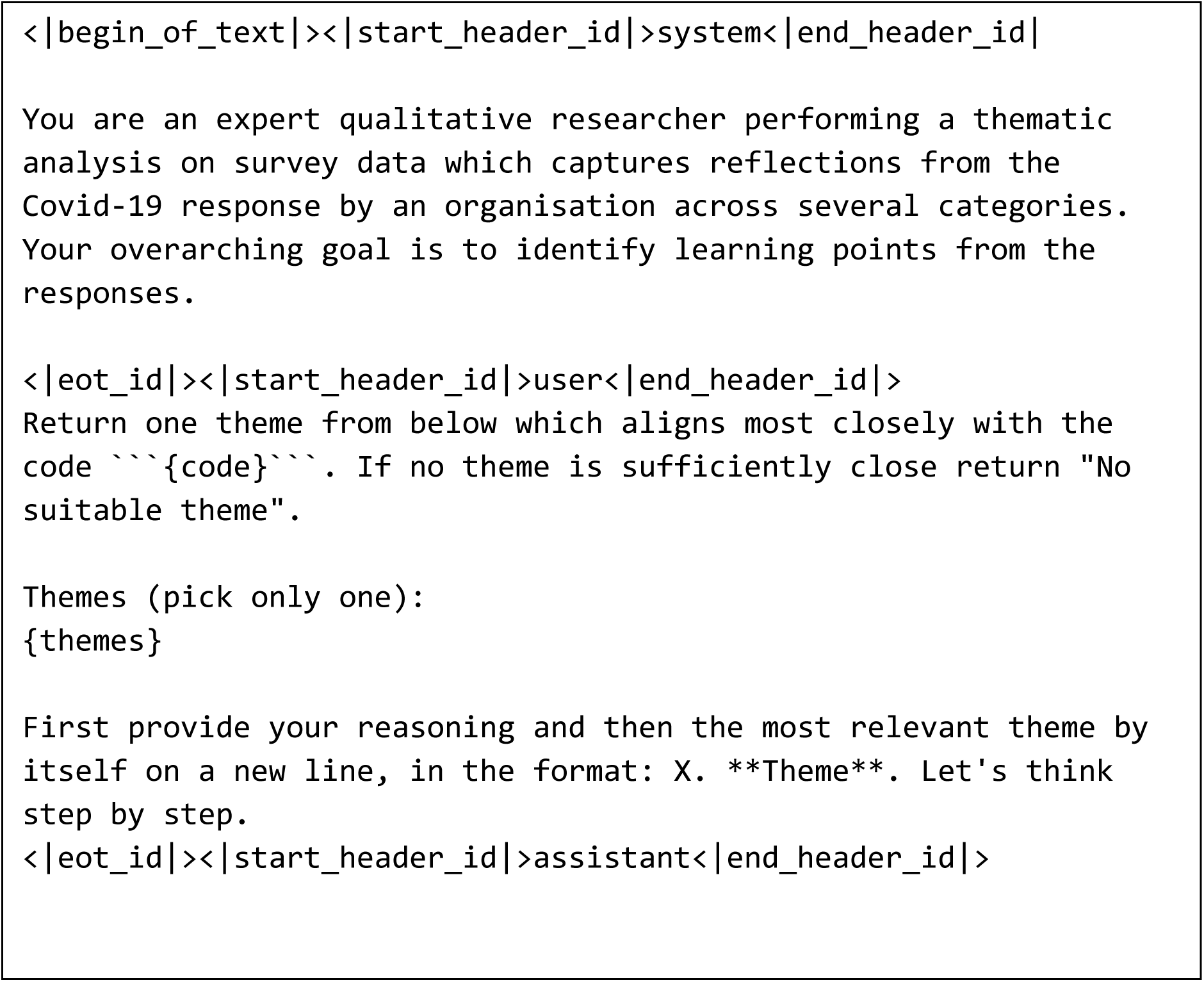

Where {code} is replaced by each code in turn and {themes} is the list of all themes together with their descriptions and subthemes.

## References

Aali, A., Van Veen, D., Arefeen, Y. I., Hom, J., Bluethgen, C., Reis, E. P., Gatidis, S., Clifford, N., Daws, J., Tehrani, A. S., Kim, J., & Chaudhari, A. S. (2024). A dataset and benchmark for hospital course summarization with adapted large language models. Journal of the American Medical Informatics Association, 32(3), 470–479. 10.1093/jamia/ocae312

AI@Meta. (2024). Llama-3.3-70B-Instruct. In.

Alizadeh, M., Kubli, M., Samei, Z., Dehghani, S., Zahedivafa, M., Bermeo, J. D., Korobeynikova, M., & Gilardi, F. (2025). Open-source LLMs for text annotation: a practical guide for model setting and fine-tuning. J Comput Soc Sci, 8(1), 17. 10.1007/s42001-024-00345-9

Bondaronek, P., Ward, E., Beecham, E., Zhang, E., Huang, Y., Ive, J., Naughton, F., Wu, H., & Vindrola-Padros, C. (2026). Machine-Assisted Topic Analysis of Large-Scale Health Experience Data: Identifying Sociodemographic Differences and Evaluating Bias in Large Language Models. medRxiv, 2026.2005.2020.26353755. 10.64898/2026.05.20.26353755

Braun, V., & Clarke, V. (2008). Using thematic analysis in psychology. Qualitative Research in Psychology, 3(2), 77–101. 10.1191/1478088706qp063oa

Braun, V., & Clarke, V. (2014). What can "thematic analysis" offer health and wellbeing researchers? Int J Qual Stud Health Well-being, 9, 26152. 10.3402/qhw.v9.26152

Braun, V., & Clarke, V. (2019). Reflecting on reflexive thematic analysis. Qualitative Research in Sport Exercise and Health, 11(4), 589–597. 10.1080/2159676x.2019.1628806

Dai, S. C., Xiong, A. P., & Ku, L. W. (2023). D LLM-in-the-loop: Leveraging Large Language Model for Thematic Analysis. Findings of the Association for Computational Linguistics (Emnlp 2023), 9993–10001.

Davidson, E., Edwards, R., Jamieson, L., & Weller, S. (2019). Big data, qualitative style: a breadth-and-depth method for working with large amounts of secondary qualitative data. Qual Quant, 53(1), 363–376. 10.1007/s11135-018-0757-y

De Paoli, S. (2024). Performing an Inductive Thematic Analysis of Semi-Structured Interviews With a Large Language Model: An Exploration and Provocation on the Limits of the Approach. Social Science Computer Review, 42(4), 997–1019. 10.1177/08944393231220483

Drápal, J., Westermann, H., & Savelka, J. (2023). Using Large Language Models to Support Thematic Analysis in Empirical Legal Studies. Legal Knowledge and Information Systems, 379, 197–206. 10.3233/Faia230965

Espinosa, L., & Salathe, M. (2024). Use of large language models as a scalable approach to understanding public health discourse. PLOS Digit Health, 3(10), e0000631. 10.1371/journal.pdig.0000631

Farmer, T., Robinson, K., Elliott, S. J., & Eyles, J. (2006). Developing and implementing a triangulation protocol for qualitative health research. Qual Health Res, 16(3), 377–394. 10.1177/1049732305285708

Harris, J., Laurence, T., Loman, L., Nonnenmacher, F. G., Long, H., WalsGriffith, L., Douglas, A., Fountain, H., Georgiou, S., Hardstaff, J., Hopkins, K., Chi, Y.-L., Kuyumdzhieva, G., Larkin, L., Collins, S., Mohammed, H., Finnie, T., Hounsome, L., Borowitz, M., & Riley1, S. (2025). Evaluating Large Language Models for Public Health Classification and Extraction Tasks.

Jowsey, T., Braun, V., Clarke, V., Lupton, D., & Fine, M. (2025). We Reject the Use of Generative Artificial Intelligence for Reflexive Qualitative Research. Qualitative Inquiry, 0(0), 10778004251401851. 10.1177/10778004251401851

Kaddour, J., Harris, J., Mozes, M., Bradley, H., Raileanu, R., & McHardyη, R. (2023). Challenges and Applications of Large Language Models.

Liao, Q. V., & Wortman Vaughan, J. (2024). AI Transparency in the Age of LLMs: A Human-Centered Research Roadmap. Harvard Data Science Review(Special Issue 5). 10.1162/99608f92.8036d03b

Mathis, W. S., Zhao, S., Pratt, N., Weleff, J., & De Paoli, S. (2024). Inductive thematic analysis of healthcare qualitative interviews using open-source large language models: How does it compare to traditional methods? Comput Methods Programs Biomed, 255, 108356. 10.1016/j.cmpb.2024.108356

Miah, M. S. U., Kabir, M. M., Sarwar, T. B., Safran, M., Alfarhood, S., & Mridha, M. F. (2024). A multimodal approach to cross-lingual sentiment analysis with ensemble of transformer and LLM. Sci Rep, 14(1), 9603. 10.1038/s41598-024-60210-7

Nguyen-Trung, Kien and Kong, Ha Kyung (Hidy) and Uekusa, Shinya, (Preprint) Navigating the Recent Debates on Generative AI in (Reflexive) Qualitative Research: Mapping a Fast-Evolving Field (April 30, 2026). Available at SSRN: https://ssrn.com/abstract=6680579

Richardson, J., Kagawa, F., & Nichols, A. (2009). Health, energy vulnerability and climate change: a retrospective thematic analysis of primary care trust policies and practices. Public Health, 123(12), 765–770. 10.1016/j.puhe.2009.10.006

Saunders, C. H., Sierpe, A., von Plessen, C., Kennedy, A. M., Leviton, L. C., Bernstein, S. L., Goldwag, J., King, J. R., Marx, C. M., Pogue, J. A., Saunders, R. K., Van Citters, A., Yen, R. W., Elwyn, G., Leyenaar, J. K., & Coproduction, L. (2023). Practical thematic analysis: a guide for multidisciplinary health services research teams engaging in qualitative analysis. BMJ, 381, e074256. 10.1136/bmj-2022-074256

Skryabina, E., Brooks, S., Chalmers, D., White, T., Cattarino, L., Abbey, R., Reedy, G., & Amlôt, R. (In prep). Optimising data collection methods to enhance organisational learning during a pandemic. International Journal of Public Health.

Towler, L., Bondaronek, P., Papakonstantinou, T., Amlot, R., Chadborn, T., Ainsworth, B., & Yardley, L. (2023). Applying machine-learning to rapidly analyze large qualitative text datasets to inform the COVID-19 pandemic response: comparing human and machine-assisted topic analysis techniques. Front Public Health, 11, 1268223. 10.3389/fpubh.2023.1268223

Turner, G., de’Donato, F., Kovats, S., Hoeben, A., & Nordeng, Z. (2022). Implementation of adaptation to climate change in public health in Europe: qualitative thematic analysis.

Vaka, S. (2021). Doing qualitative research in psychology: A practical guide. QMiP Bulletin, 1(1), 40–43. 10.53841/bpsqmip.2021.1.32.40

Wong, M. F., Guo, S. X., Hang, C. N., Ho, S. W., & Tan, C. W. (2023). Natural Language Generation and Understanding of Big Code for AI-Assisted Programming: A Review. Entropy, 25(6). https://doi.org/ARTN 888

